# Report on COVID-19 Verification Case Study in Nine Countries Using the SIQR model

**DOI:** 10.1101/2020.10.07.20208298

**Authors:** Reiji Suda

## Abstract

This report uses the SIQR model proposed by Takashi Odagaki to examine the epidemic trend of COVID-19 in nine major countries during February-May 2020, and to clarify the peculiar trend of infection in Japan. The SIQR model, which is an improvement on the conventional SIR model, is unique in that it allows us to theoretically clarify the epidemic phenomenon by separating the number of daily confirmed new cases by testing and the number of infecteds at large who remain untested, and also allows us to theoretically consider measures to control the epidemic. The infection control measures of each country were analyzed by dividing them into three groups according to the size of the decay (or growth) rate of infected at large (λ). The active group includes China and South Korea, the passive group includes the United States and Sweden, and the average group includes Germany, Italy, France, Spain, and Japan. China and South Korea are the countries with the best testing and quarantine systems, and South Korea in particular having managed to contain the infection without lockdown through early quarantine by thorough testing. On the other hand, the United States and Sweden do not have a well-developed inspection and quarantine system and have shown little restraint in social distancing. In the case of Japan, the following special factors may have contributed to the extreme lack of PCR testing : (1) The “4-day fever rule” established by the Ministry of Health, Labour and Welfare was strictly enforced. (2) Even after the decision to postpone the Olympics, the government continued to monopolize PCR testing for the sake of unified analysis of infection data, and the policy of expanding PCR testing by private companies was not implemented.

## 1. Introduction

Using the SIQR model proposed by Takashi Odagaki^1)^, this report examines the epidemic trend of COVID-19 from February to May 2020 in nine major countries, and identifies specific trends in the actual state of infection in Japan.

When the Diamond Princess returned to the port of Yokohama on February 3, the total number of infected people on board was 712. While I was keeping a close eye on the situation after the ship began disembarking on February 19, I noticed that the trend in the number of infected people in Japan during March and April differed from the rest of the world.

Figure 1 compares the cumulative number of people infected after reaching 100 by country, and it can be seen that Japan’s trend is quite pequliar.

**Fig.1.**
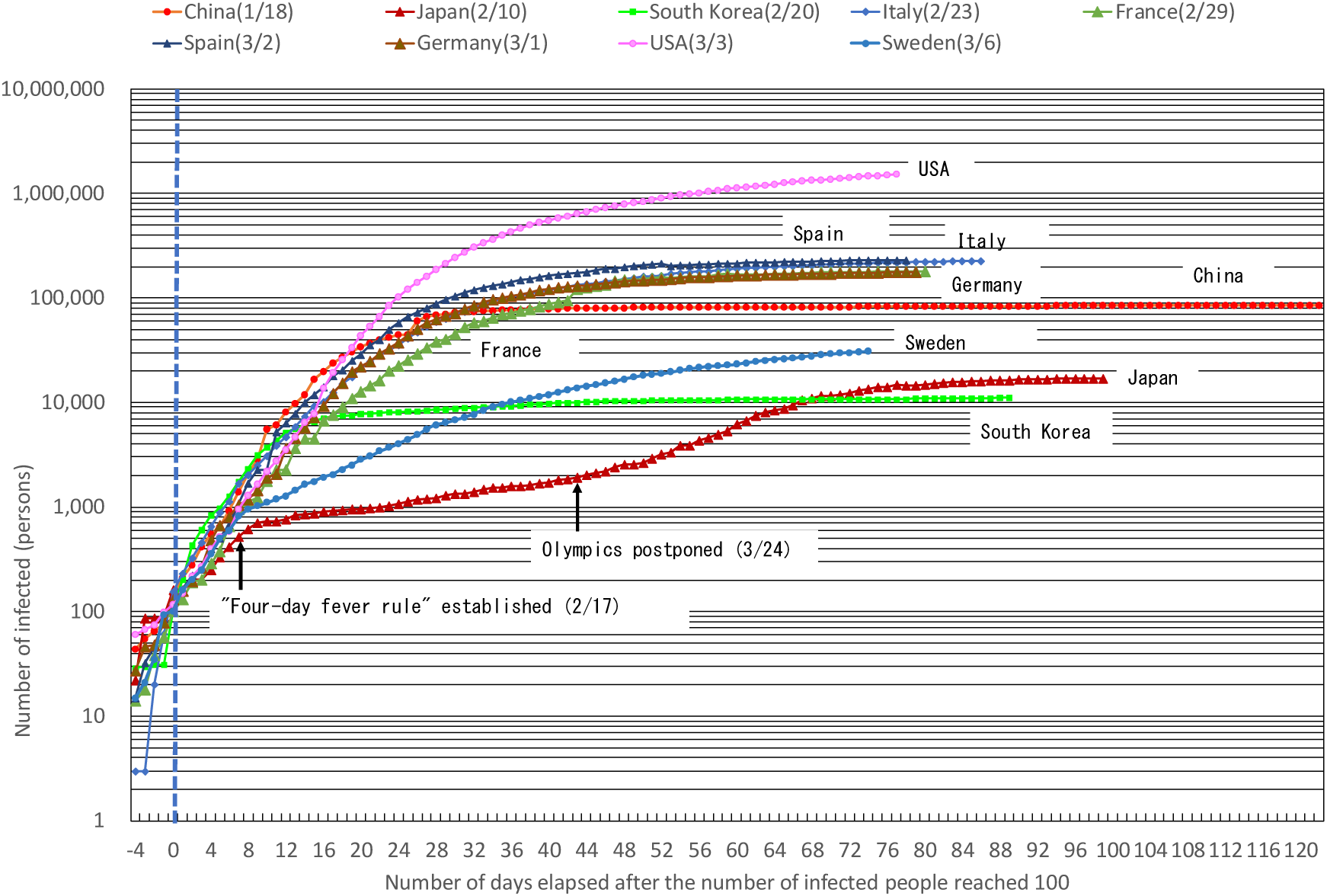
Comparison of the number of people infected after reaching 100 by country. Note 1: In the legend, countries in parentheses indicate the month and date of reaching 100 cases. Note 2: Data on the number of infected people are through May 19. Note 3: The number of infected people in Japan includes the Diamond Princess.

Why are Japan’s trends up to the date of the decision to postpone the Olympics (3/24) different from those of Germany and France? Why is this different from Sweden, which also adopted the herd immunity policy? And why is the number of cases of infection increasing rapidly after the date of the decision to postpone? Through theoretical analysis of these questions using the SIQR model, we summarize below how the situation was progressing.

## 2. Methods of Analysis

First, we summarize the method of the SIQR model based on the paper by Takashi Odagaki. The SIQR model, which is a modification of the conventional SIR model, is unique in that it provides a theoretical understanding of epidemic phenomena by separating quarantined infected people identified by testing and community-acquired people who remain untested, and also provides a theoretical consideration of measures to control the epidemic.

### 2.1 Setting the Basic Formula

The statistical data reported daily is the “number of new infections”, which can be treated as the “number of positive cases per day” and can be expressed as ΔQ(t) = qI(t) in the SIQR model.

That is, I(t) = ΔQ(t)/q = ΔQ(t)/(*β*N - *γ* - λ). where I(t) is the number of infecteds at large, (q) is the quarantine rate, *β*N is the transmission coefficient, and *γ* is the removal rate and λ are determinants of the decay (or growth) rate of infected at large, which can be expressed as (*β*N-q-*γ*).

The next step is to determine the effectiveness of social distancing (a) and the enhancement factor of quarantine rate (b), which is a factor of human-induced infection control measures, as well as the rate of increase or decrease in the number of new define λ = (1-q’-a)*β*N-qb-*γ* as the λ that takes into account the quarantine rate of the new patients immediately after they are infected (q’) of infection-positive individuals to be.

Since no one is isolated immediately after infection due to a long incubation period, the quarantine rate of the new patients immediately after they are infected (q’) shall be regarded as zero in principle.

### 2.2 Verification Procedure

The “number of positive cases per day” ΔQ(t) is approximated by an exponential function ΔQ(to)e^λ(t-to)^ with a coefficient of λ, as in the case of the number of infecteds large I(t). Based on the distribution of the number of positive cases per day, we also classify them into an initial phase, an expansion phase, a transition phase, and a decay phase, and establish a continuous period classification, from the first to the fourth period. The coefficient λ and the initial value ΔQ(to) are determined by obtaining an exponential approximation curve for each period based on the actual values. In the case of China and Korea, the period of spread of infection is earlier than the first period and is considered to be the expansion phase. Next, *γ, β*N, q, a, and b are set as follows.

1. Since the removal rate *γ* of community-infected persons is considered to be almost constant during the entire period, the number of days of cure is estimated to be 33 days, and the inverse of 0.03 is set.
2. The quarantine rate q for the 2nd period (expansion phase) is considered to be very low, and so set q = 0.
3. The transmission coefficient *β*N in the 2nd period (expansion phase) are set based on the coefficient λ determined by an exponential approximation curve using data from the 2nd period(*β*N=λ+q+*γ*). This *β*N is used in the 1st, 3rd and 4th period.
4. The quarantine rate q in the 3rd period (transition phase) is determined by an exponential approximation curve based on the 3rd period data. It is set up from the coefficient λ (as close to 0 as possible), *β*N and *γ*. (q = *β*N-*γ*-λ). The same q is used in the 1st and 4th period. 5) The effectiveness of social distancing (a) and the enhancement factor of quarantine rate (b) in the 4th (decay) period should be set so that λ = (1-q’-a) *β*N-qb-*γ* based on the factor λ determined by the exponential approximation curve based on the 4th period data and the previously set *β*N, q, and *γ*. The λ re-set after setting (a) and (b) is defined as the amount for determining the decay (or growth) rate of infected at large. The default value of the effectiveness of social distancing (a) is 0, and the default value of the enhancement factor of quarantine rate (b) is 1.0. In the 1st (initial) period, (b) is treated as variable, and in the 4th (decay) period, (a)and (b) are treated as variable.

## 3. Analysis of Country Data

Based on the above analysis, we will analyze the actual number of infected people in Japan and Tokyo, as well as the actual number of infected people in other countries, to clarify the situation in each country and the parameters of the SIQR model.

### 3.1 Countries and data used for verification

In addition to Japan and Tokyo, the countries selected as COVID-19 verification cases using the SIQR model are those with distinctive infection control measures, in the order of the earliest date of the cumulative number of infected people reaching 100. The eight countries are China (1/18), South Korea (2/20), Italy (2/23), France (2/29), Germany (3/1), Spain (3/2), the United States (3/3), and Sweden (3/6). The figures in parentheses indicate the month and date when the cumulative number of cases reached 100, and that in Japan is 2/10.

For the graph of theoretical estimates of the number of infecteds at large and confirmed new cases, it is assumed that the confirmed new cases occurred two weeks after the community infection at large and that the confirmed new cases occurred one week later in South Korea due to the early response.

The data for Japan used in the theoretical analysis are from the Toyo Keizai ONLINE data^2)^, which were published by the Ministry of Health, Labour and Welfare. For the purpose of analysis, data from the Diamond Princess were excluded. Data for Tokyo are from the Tokyo Metropolitan Government^3)^ and data for other countries are from Johns Hopkins University^4)^. Data on population and number of tests conducted are from Worldometers’s COVID-19 ^5)^.

### 3.2 Verification using Japan and Tokyo data

Due to the peculiarity of Japan’s trend up to the date of the decision to postpone the Olympics (3/24), the 1st period is set for 2/18 to 3/23. The 2nd expansion period is set for 3/24 to 4/10, the 3rd transition period is set for 4/11 to 4/18, and the 4th decay period is set for 4/19 to 5/19. The theoretical analysis and parameter verification values based on actual daily positive case data are summarized in Table 1, together with those of foreign countries.

**Table 1.**
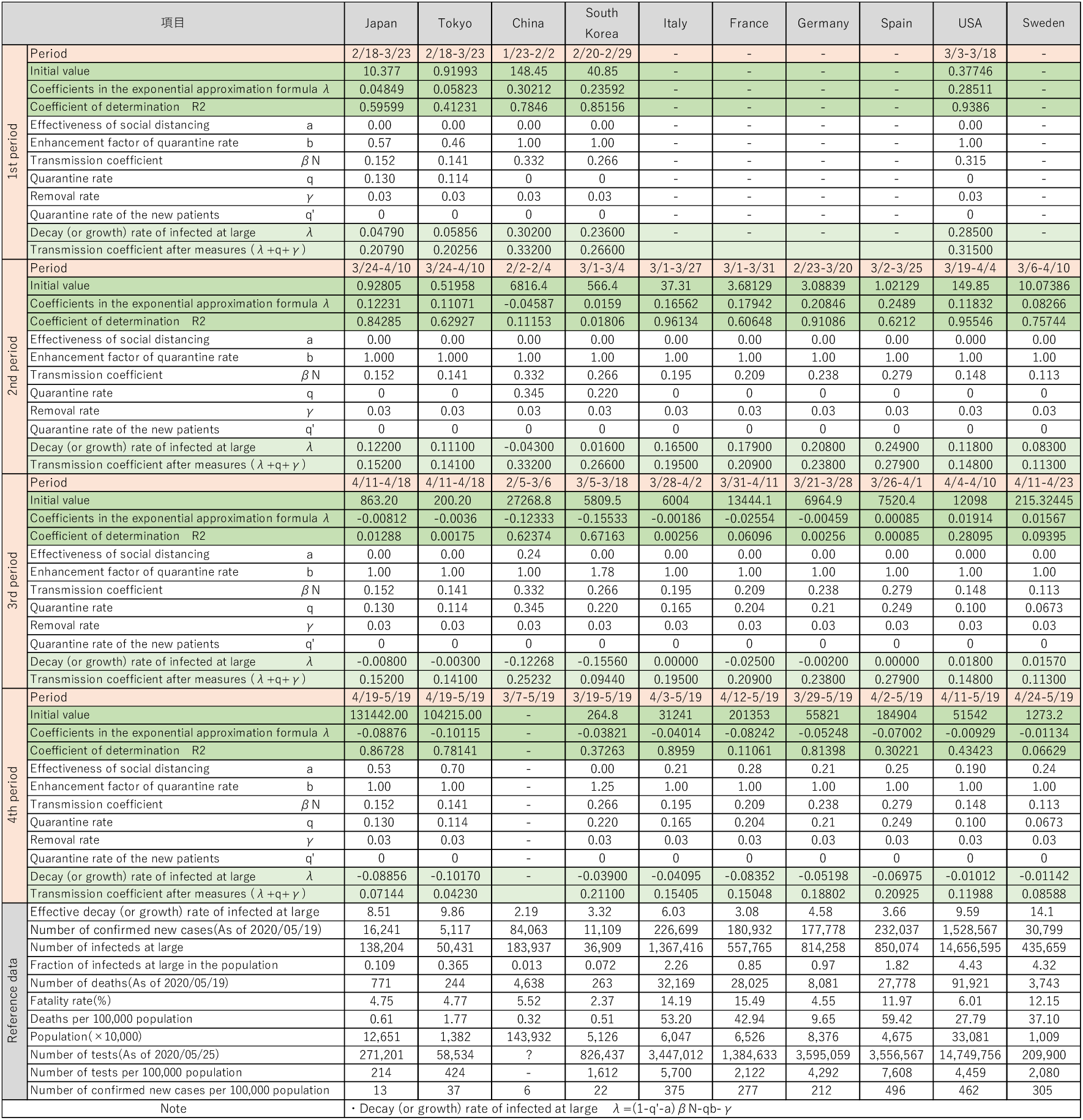
Theoretical analysis and Parameter verification values.

#### (1) The enhancement factor of quarantine rate (b)

In the 1st period in Japan (the inspection suppression phase), the enhancement factor of quarantine rate (b) is calculated to be 0.57 due to the λ obtained by the exponential approximation curve and the *β*N, q and *γ* determined in from the 2nd period to the 4th period, since the effectiveness of social distancing (a) can be regarded as zero. In other words, until the date of the decision to postpone the Olympic Games in Japan (3/24), Japan had taken about 60% of the measures to prevent the isolation of people from the second and subsequent phases by suppressing testing.

#### (2) The effectiveness of social distancing (a)

In the 4th decay period in Japan, the effectiveness of social distancing (a) is calculated to be 0.53 due to the λ obtained by the exponential approximation curve and the *β*N, q, and *γ* determined in the 2nd and 3rd period, since the enhancement factor of quarantine rate (b) can be regarded as 1.0. This means that the behavioral restraint rate during the decay phase in Japan is estimated to have been about 50%.

Using the same method, the effectiveness of social distancing (a) during the decay phase based on Tokyo data is 0.70, and it is estimated that about 70% of people in Tokyo refrained from contacting each other.

#### (3) Transmission coefficient *β*N and Quarantine rate of infecteds at large (q)

The transmission coefficient *β*N in Japan is 0.152, and the quarantine rate of infecteds at large (q) is 0.13. The results are quite small in comparison to that of other countries mentioned below.

#### (4) Fraction of infecteds at large in the population

The effective decay (or growth) rate of infected at large in Japan is 8.52, and for every 16,000 positive cases, there are approximately 140,000 infecteds at large, with the fraction of infecteds at large in the population of 0.109%.

According to the results of the verification of Tokyo data using the same method, the effective decay (or growth) rate of infected at large was 9.86. It is estimated that there are approximately 50,000 infecteds at large compared to 5,000 positive cases, with the fraction of infecteds at large in the population of 0.365%.

Figures 2 and 3 show the graphs of confirmed new cases and exponential approximation curves for Japan and Tokyo. In addition, assuming that confirmed new cases occur 2 weeks after community infection, the graphs of the estimation of the number of infecteds at large and confirmed new cases in Japan and Tokyo are shown in Figures 4 and 5.

**Fig.2.**
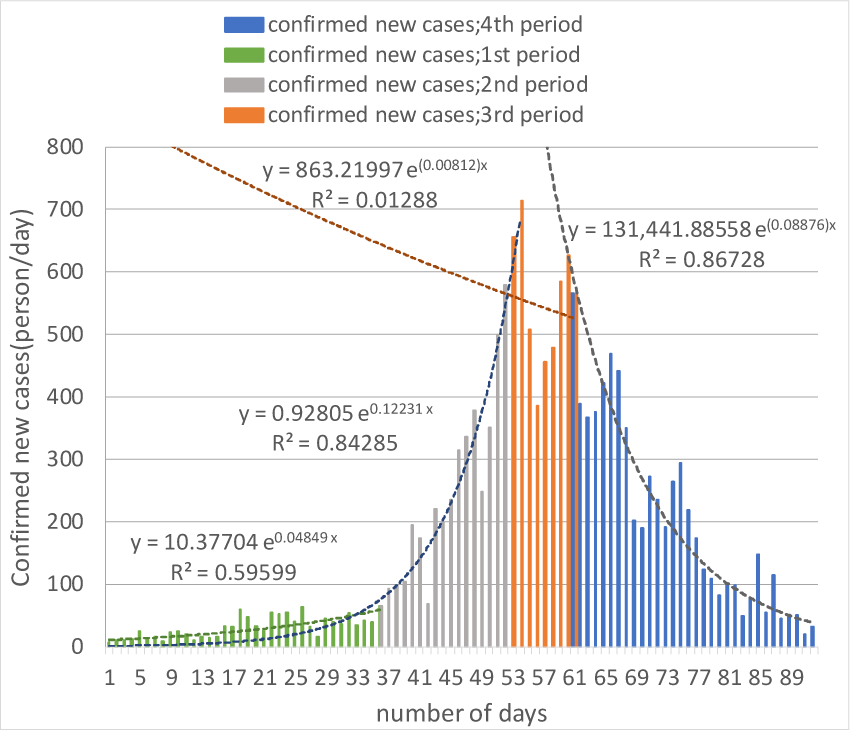
Confirmed new cases and Exponential approximation curve (Japan)

**Fig.3.**
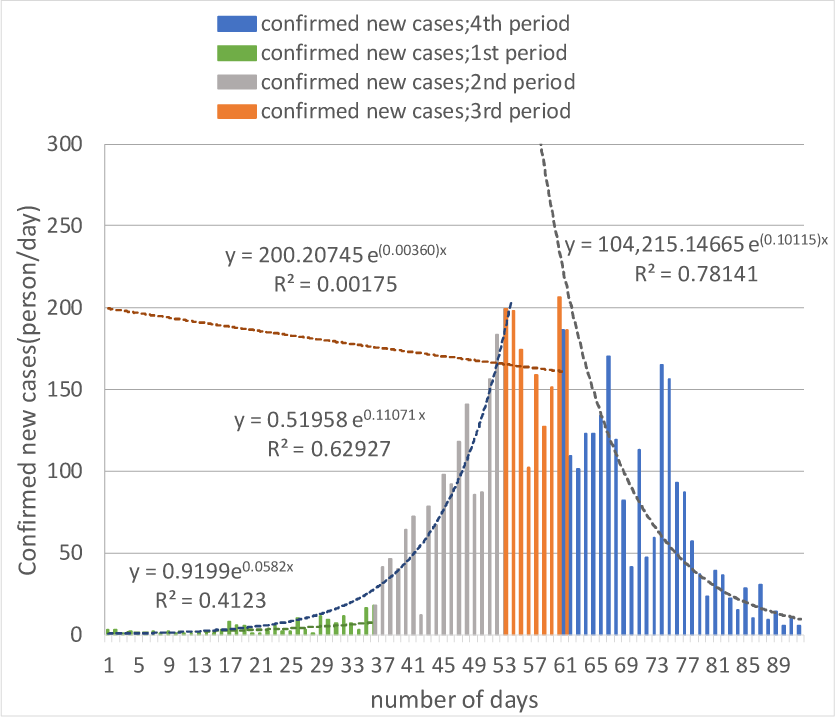
Confirmed new cases and Exponential approximation curve (Tokyo)

**Fig.4.**
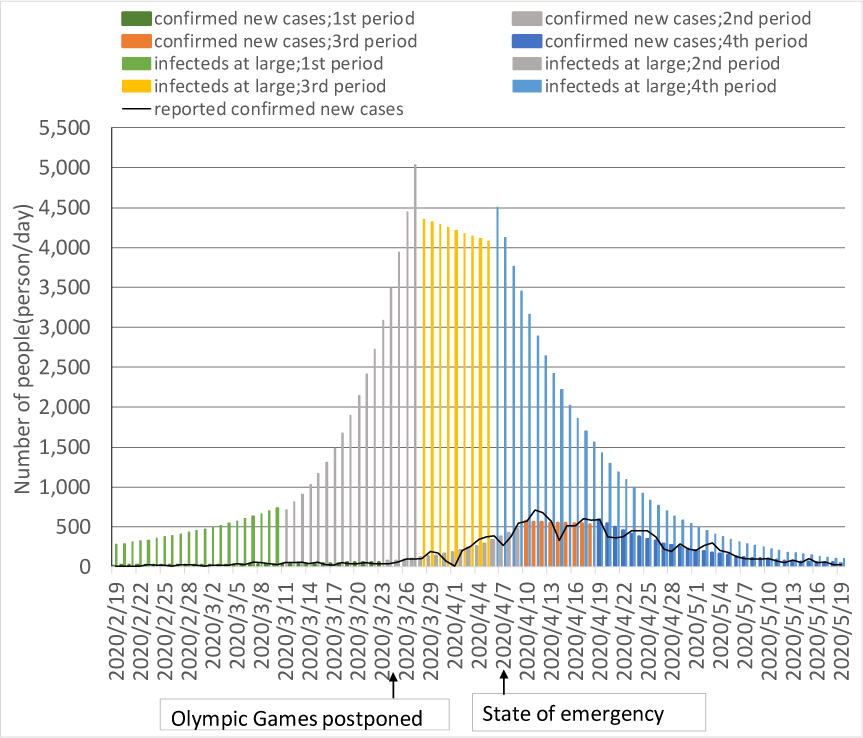
Estimation of the number of infecteds at large and confirmed new cases (Japan)

**Fig.5.**
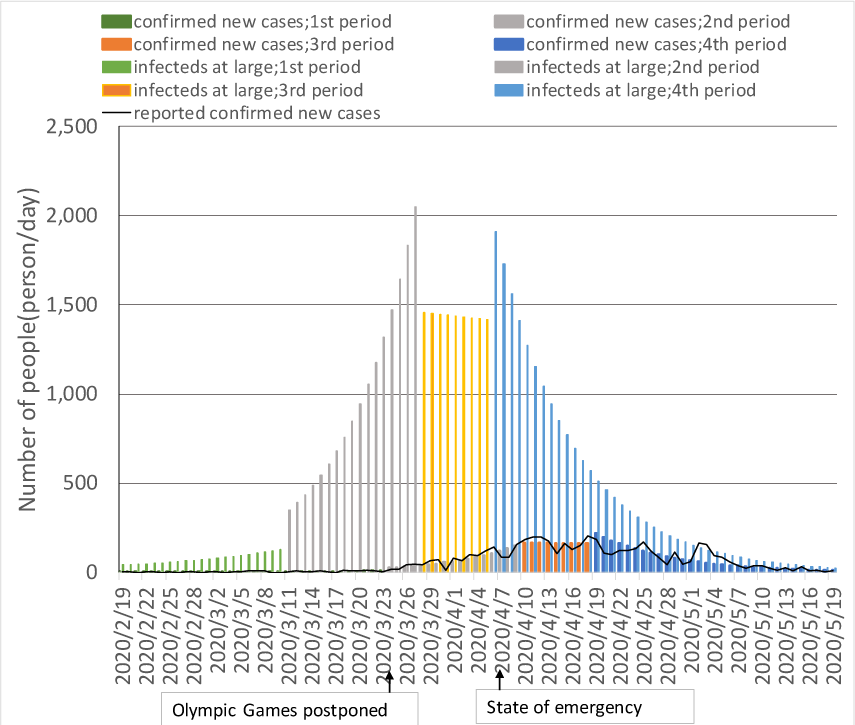
Estimation of the number of infecteds at large and confirmed new cases (Tokyo)

Figure 4 shows that the peak of infection in the city is around April 1, the middle of the transition period, and the state of emergency is declared six days later in April. It shows that it was issued on the 7th. Also, the date of the decision to postpone the Olympics (3/24) would have been in the middle of a citywide outbreak.

### 3.3 Verification using foreign data

#### (1) Verification using China data

The period classification in China is set as follows: the first expansion period is set for 1/23-2/2, the second transition period is set for 2/2-2/4, and the third decay period is set for 2/5-3/6. The transmission coefficient was 0.332, the highest among the nine countries, and the quarantine rate of community-infected persons was 0.345, which is the highest among the nine countries. The effective decay (or growth) rate of infected at large was 2.19, with approximately 180,000 infecteds at large compared to 83,000 positive cases. The fraction of infecteds at large in the population is estimated to be 0.013% (see Figures 6, 7 and Table 1).

**Fig.6.**
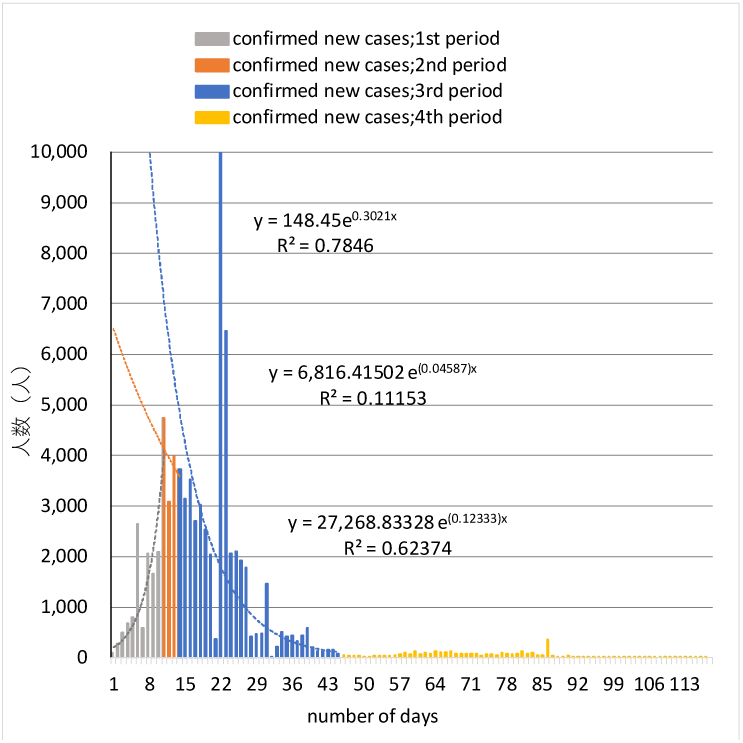
Confirmed new cases and Exponential approximation curve (China)

**Fig.7.**
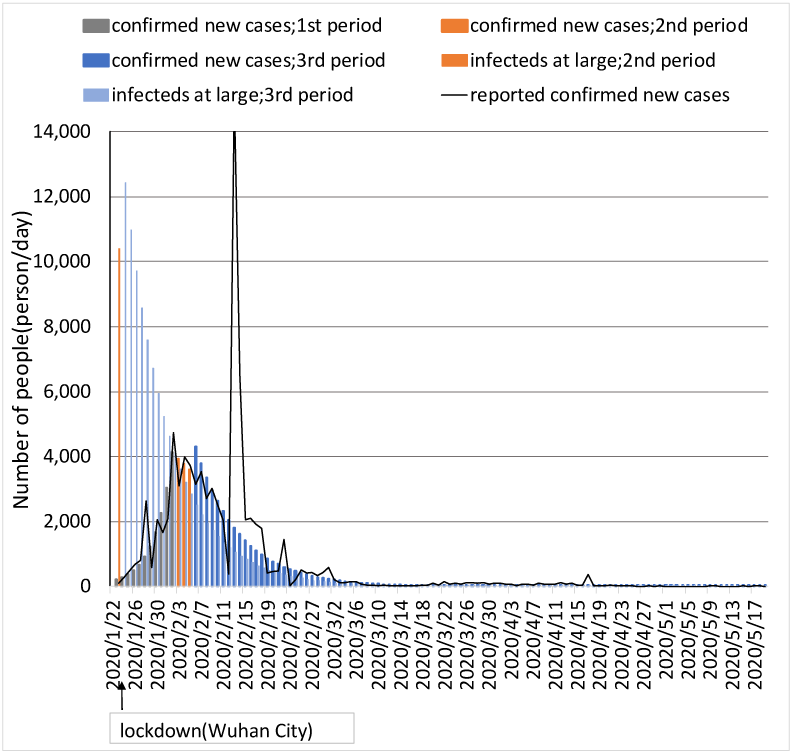
Estimation of the number of infecteds at large and confirmed new cases (China)

**Fig.8.**
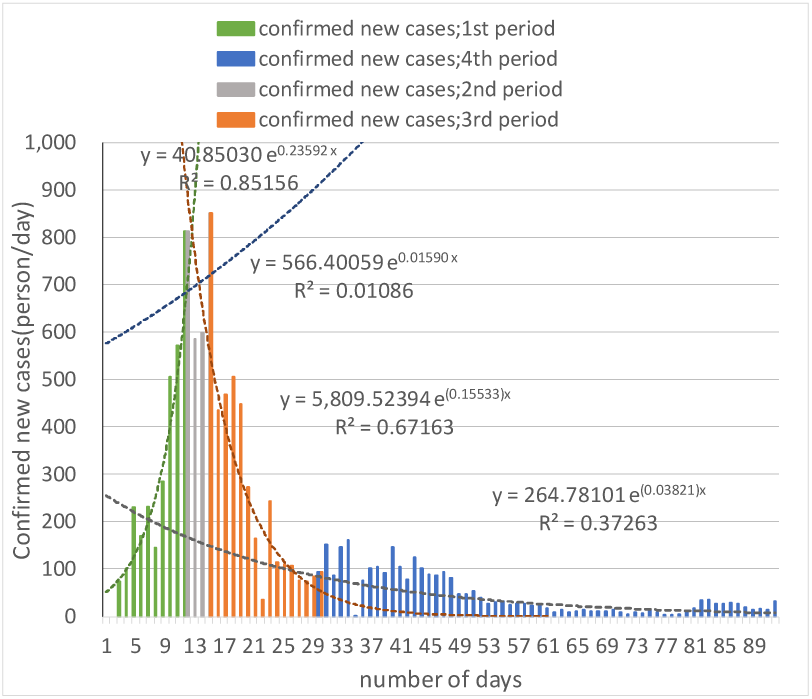
Confirmed new cases and Exponential approximation curve (South Korea)

**Fig.9.**
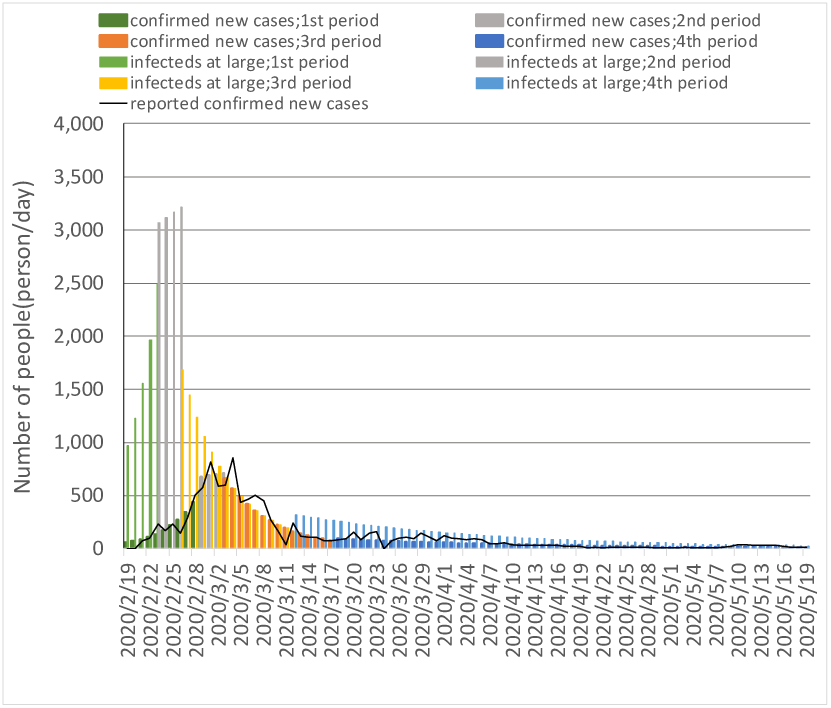
Estimation of the number of infecteds at large and confirmed new cases (South Korea)

#### (2) Verification using South Korea data

In Korea, the 1st expansion period is set for 2/20-2/29, the 2nd transition period is set for 3/1-3/4, the 3rd decay period(phase1) is set for 3/5-3/18, and the 4th decay period(phase2) is set for 3/19-5/19. The transmission coefficient was 0.266 and the quarantine rate of community-infected persons was 0.22, which were both nearly twice as high as those in Japan. In Korea, the spread of infection was rapid and the rate of decay was fast. The enhancement factor of quarantine rate (b) in the 3rd period was 1.78, contributing to the decrease in the transmission coefficient after measures *β*N. It can be said that the policy of early detection and isolation through thorough testing is effective. The effective decay (or growth) rate of infected at large was 3.32, and for the 11,000 positive cases, there were approximately 37,000 infecteds at large. The fraction of infecteds at large in the population is estimated to be 0.072%.

#### (3) Verification using Italy data

In Italy, the period classification is set as follows: the 2nd expansion period is set for 3/1-3/27, the 3rd transition period is set for 3/28-4/2, and the 4th decay period is set for 4/3-5/19. The transmission coefficient was 0.195, and the quarantine rate of community-infected persons was 0.165, which is about 30-40% higher than that of Japan. On the other hand, the effectiveness of social distancing in the 4th period was 0.21, which was smaller than in Japan. It appears that the behavioral restraint was not as thorough as in Japan. The effective decay (or growth) rate of infected at large was 6.03, with approximately 1.37 million infecteds at large compared to 230,000 positive cases. The fraction of infecteds at large in the population is estimated to be 2.26% (see Figure 10,11 and Table 1).

**Fig.10.**
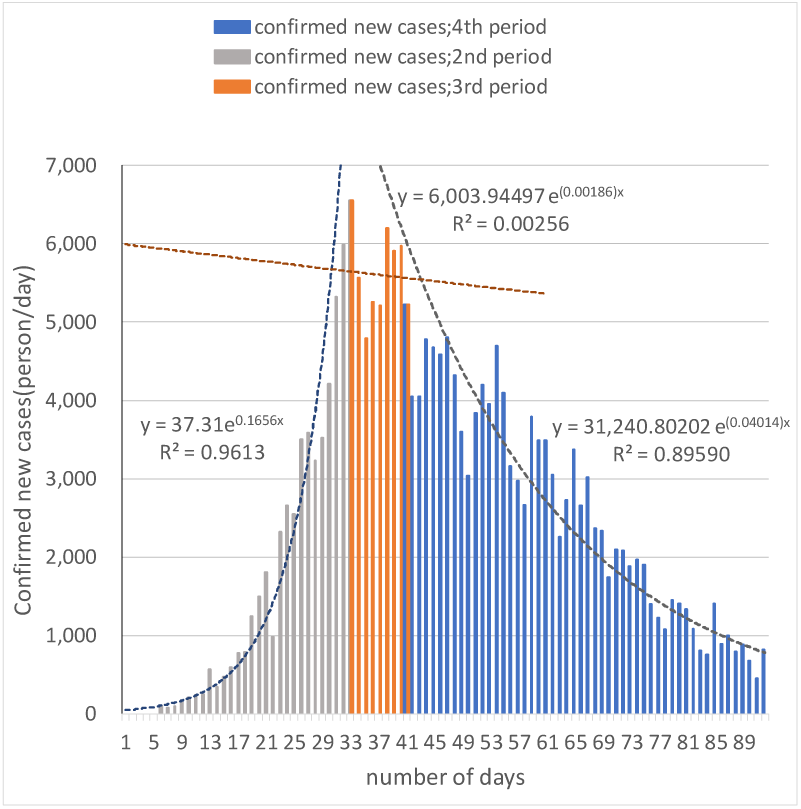
Confirmed new cases and Exponential approximation curve (Italy)

**Fig.11.**
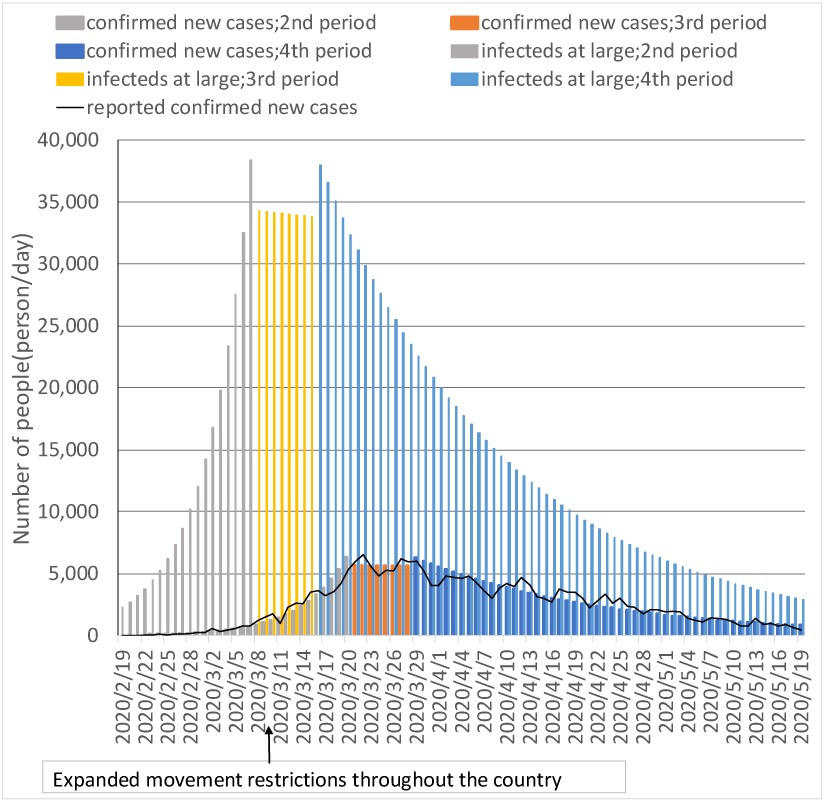
Estimation of the number of infecteds at large and confirmed new cases (Italy)

#### (4) Verification using France data

In France, the period classification is set as follows: the 2nd expansion period is set for 3/1-3/31, the 3rd transition period is set for 3/31-4/11, and the 4th decay period is set for 4/12-5/19. The transmission coefficient was 0.209, and the quarantine rate of community-infected persons was 0.204, which was 40%-50% lower than that of Japan. On the other hand, the effectiveness of social distancing in the 4th period was 0.28, which was smaller than in Japan. It appears that the behavioral restraint was not as thorough as in Japan. The effective decay (or growth) rate of infected at large was 3.08, and for every 180,000 positive cases, there are approximately 560,000 infecteds at large. The fraction of infecteds at large in the population is estimated to be 0.85% (see Figures 12,13 and Table 1).

**Fig.12.**
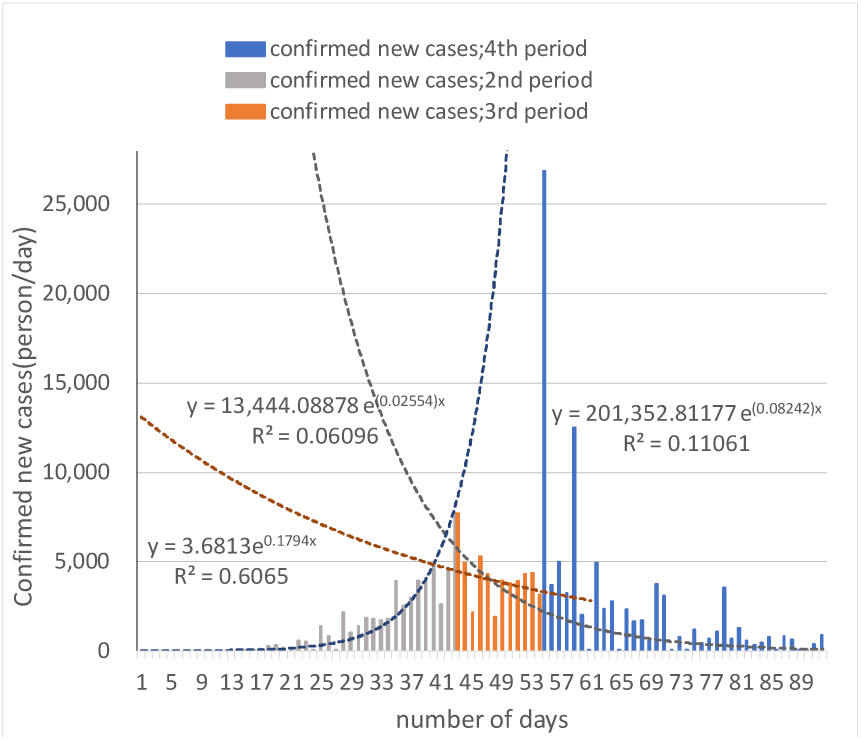
Confirmed new cases and Exponential approximation curve (France)

**Fig.13.**
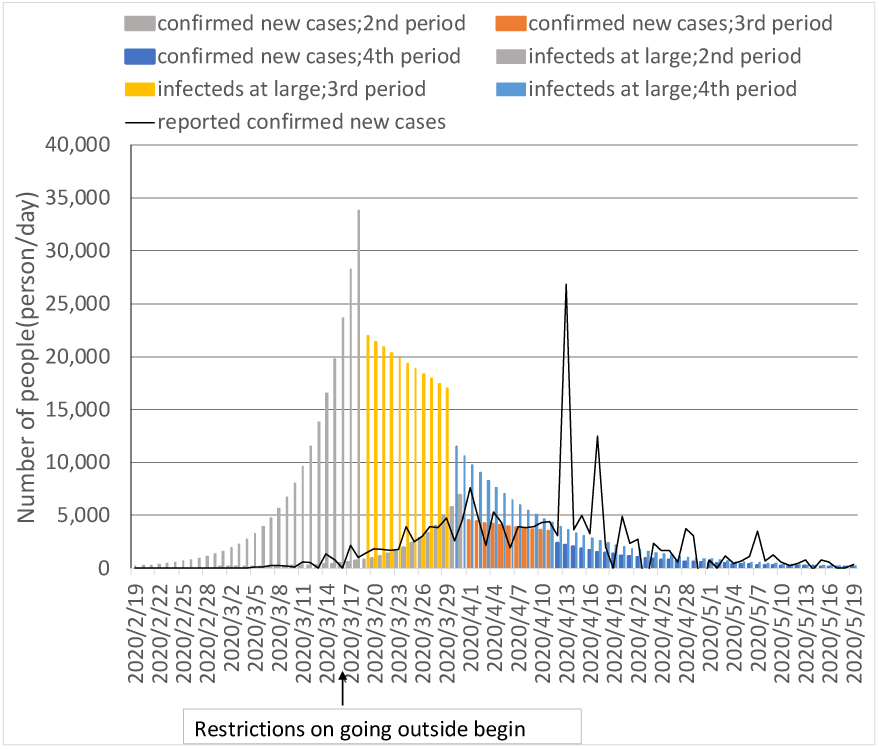
Estimation of the number of infecteds at large and confirmed new cases (France)

#### (5) Verification using Germany data

The period classification in Germany is set as follows: the 2nd expansion period is set for 2/23-3/20, the 3rd transition period is set for 3/21-3/28, and the 4th decay period is set for 3/29-5/19. The transmission coefficient was 0.238, and the quarantine rate of community-infected persons was 0.21, which was about 60%-70% higher than that of Japan. On the other hand, the effectiveness of social distancing in the 4th period was 0.21, which was smaller than in Japan. It appears that the behavioral restraint was not as thorough as in Japan. The effective decay (or growth) rate of infected at large was 4.58, with approximately 810,000 infecteds at large compared to 180,000 positive cases. The fraction of infecteds at large in the population is estimated to be 0.97% (see Figures 14,15 and Table 1).

**Fig.14.**
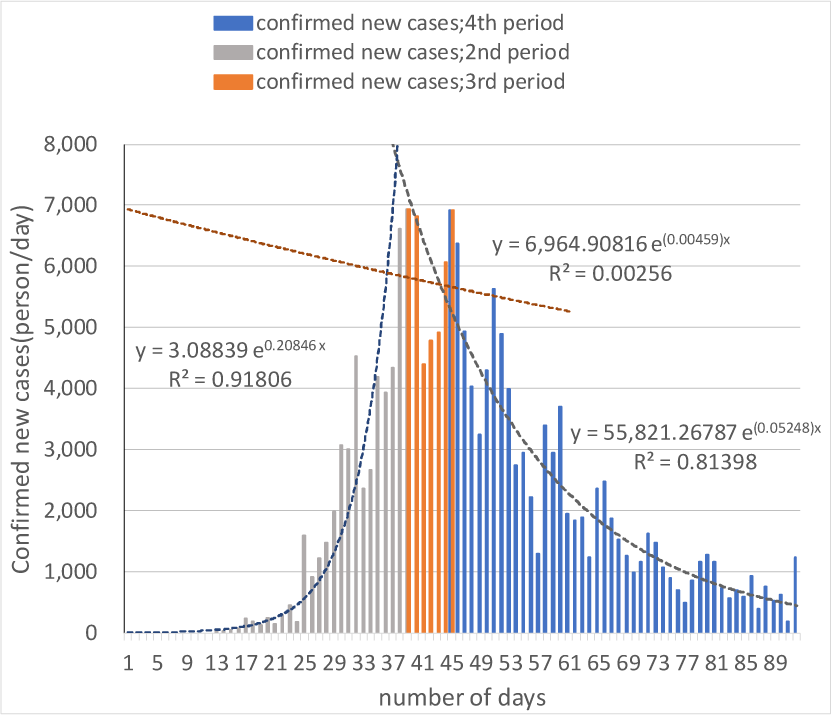
Confirmed new cases and Exponential approximation curve (Germany)

**Fig.15.**
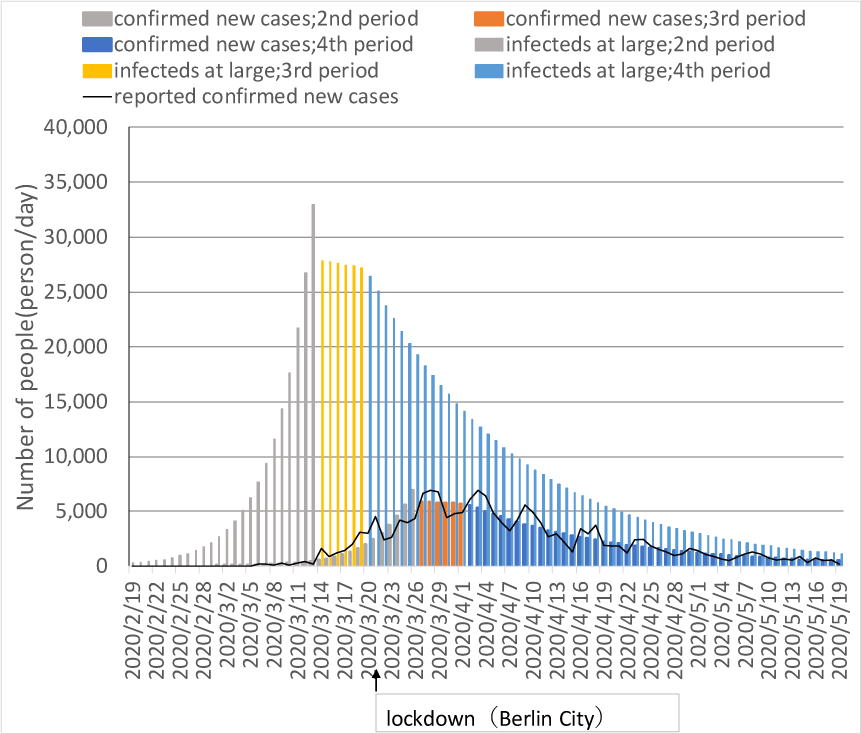
Estimation of the number of infecteds at large and confirmed new cases (Germany))

#### (6) Verification using Spain data

In Spain, the period classification is set as follows: the 2nd expansion period of is set for 3/2 to 3/25, the 3rd transition period of is set for 3/26 to 4/1, and the 4th decay period of is set for 4/2 to 5/19. The transmission coefficient was 0.279, and the quarantine rate of community-infected persons was 0.249, which was 80%-100% higher than that of Japan. On the other hand, the effectiveness of social distancing in the 4th period was 0.25, which was smaller than in Japan. It appears indicating that the behavioral restraint was not as thorough as in Japan. The effective decay (or growth) rate of infected at large was 3.66, with approximately 850,000 infecteds at large compared to 230,000 positive cases. The fraction of infecteds at large in the population is estimated to be 1.82% (see Figures 16,17 and Table 1).

**Fig.16.**
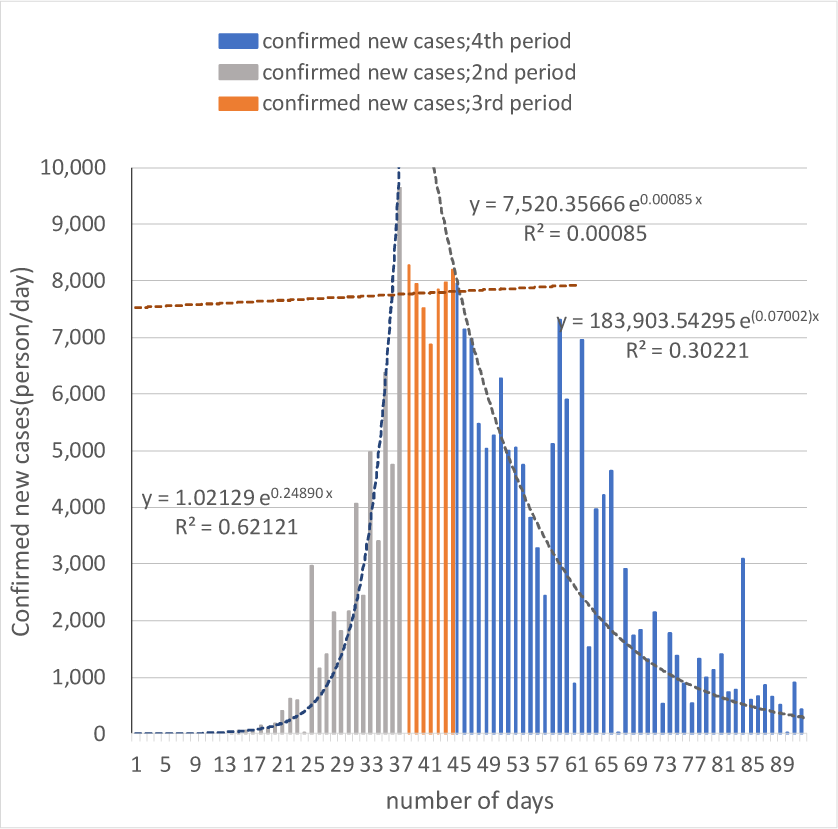
Confirmed new cases and Exponential approximation curve (Spain)

**Fig.17.**
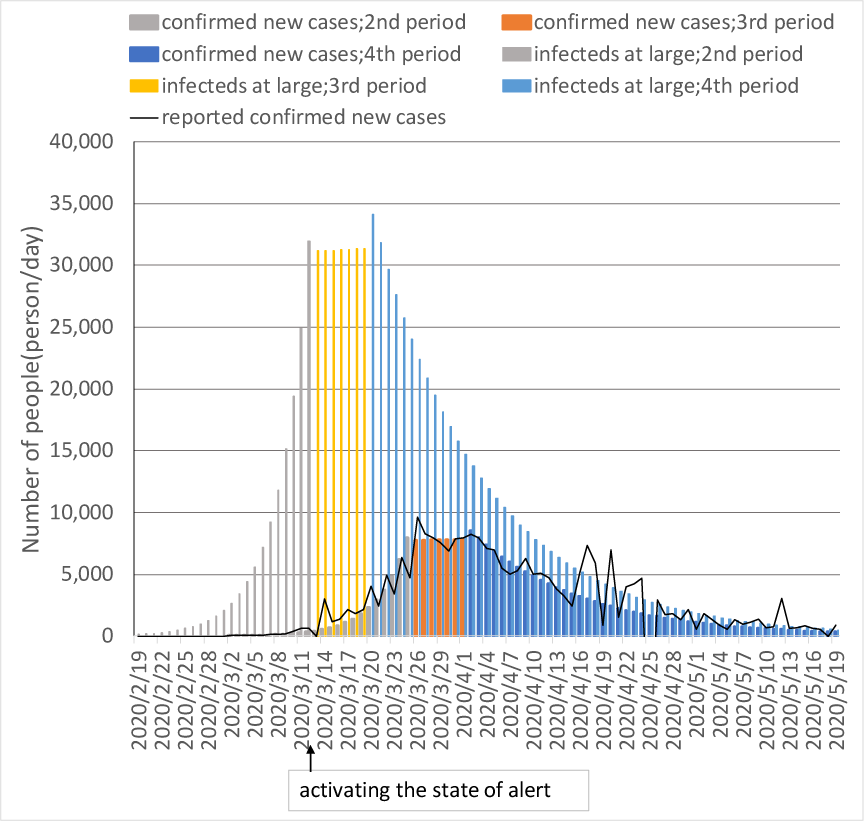
Estimation of the number of infecteds at large and confirmed new cases (Spain)

#### (7) Verification using USA data

In the U.S., the period classification is set as follows: the 1st initial period is set for 3/3 to 3/18, the 2nd expansion period is set for 3/19 to 4/4, the 3rd transition period is set for 4/4 to 4/23, and the 4th decay period is set for 4/24 to 5/19. After the 2nd period, the transmission coefficient was 0.148 and the quarantine rate of community-infected persons was 0.100, the same level as in Japan. The effective decay (or growth) rate of infected at large was 9.59, with approximately 14.65 million infecteds at large compared to 1.53 million positive cases. The fraction of infecteds at large in the population is estimated to be 4.43% (see Figures 18,19 and Table 1).

**Fig.18.**
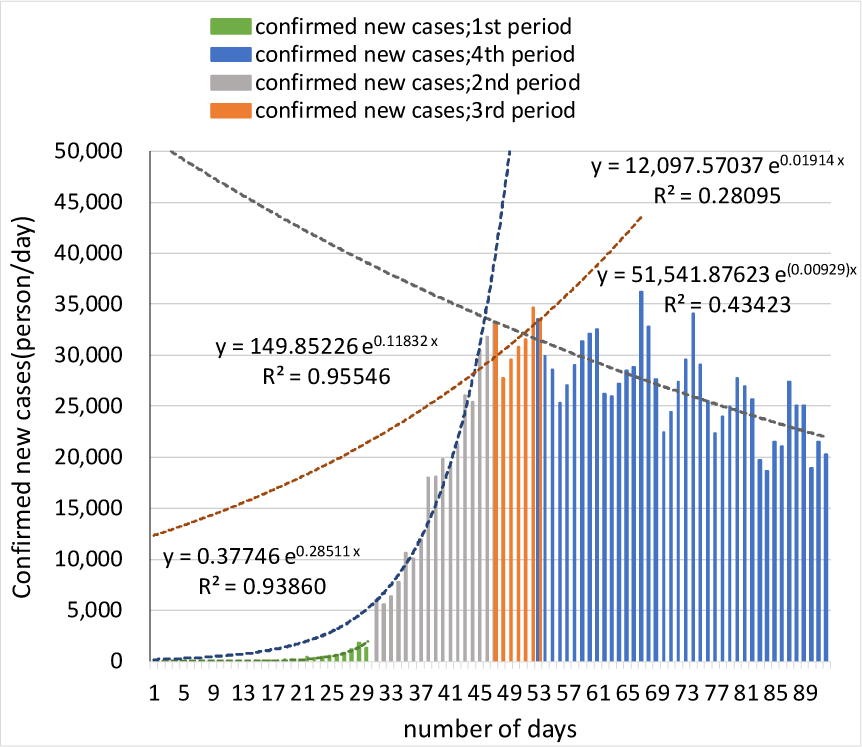
Confirmed new cases and Exponential approximation curve (USA)

**Fig.19.**
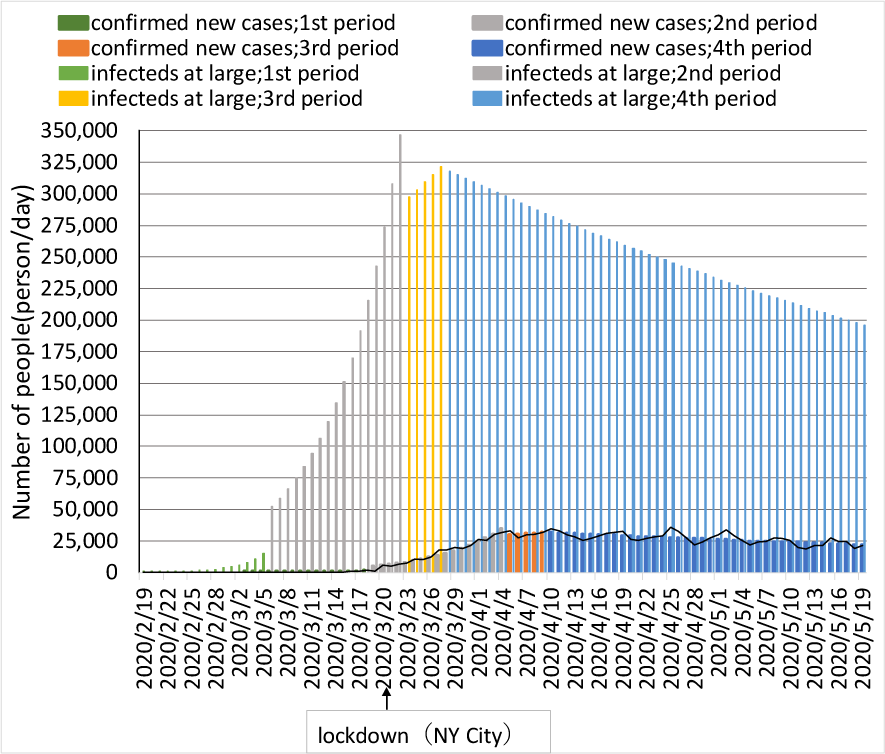
Estimation of the number of infecteds at large and confirmed new cases (USA)

#### (8) Verification using Sweden data

In Sweden, the period classification is set as follows: the 2nd expansion period is set for 3/6-4/10, the 3rd transition period is set for 4/11-4/23, and the 4th decay period is set for 4/24-5/19. The transmission coefficient was 0.113, and the quarantine rate of community-infected persons was 0.0673, which was 20%-40% lower than that of Japan. The behavioral restraint rate in the 4th decay period was 0.24, which is smaller than in Japan. It appears indicating that the behavioral restraint was not thorough. The effective decay (or growth) rate of infected at large was 14.1, with approximately 430,000 infecteds at large compared to 31,000 positive cases. The fraction of infecteds at large in the population is estimated to be 4.32% (see Figures 20,21 and Table 1).

**Fig.20.**
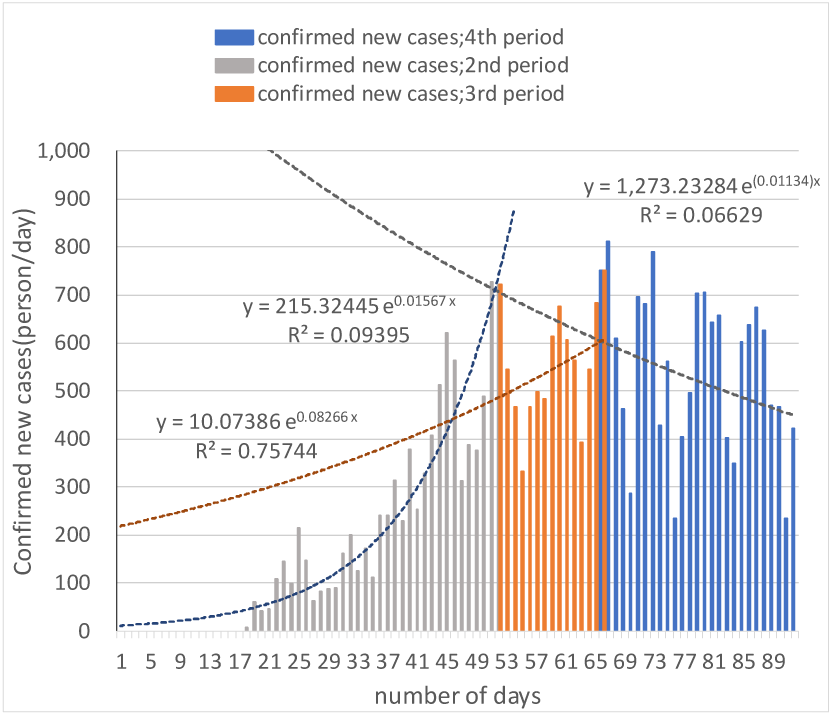
Confirmed new cases and Exponential approximation curve (Sweden)

**Fig.21.**
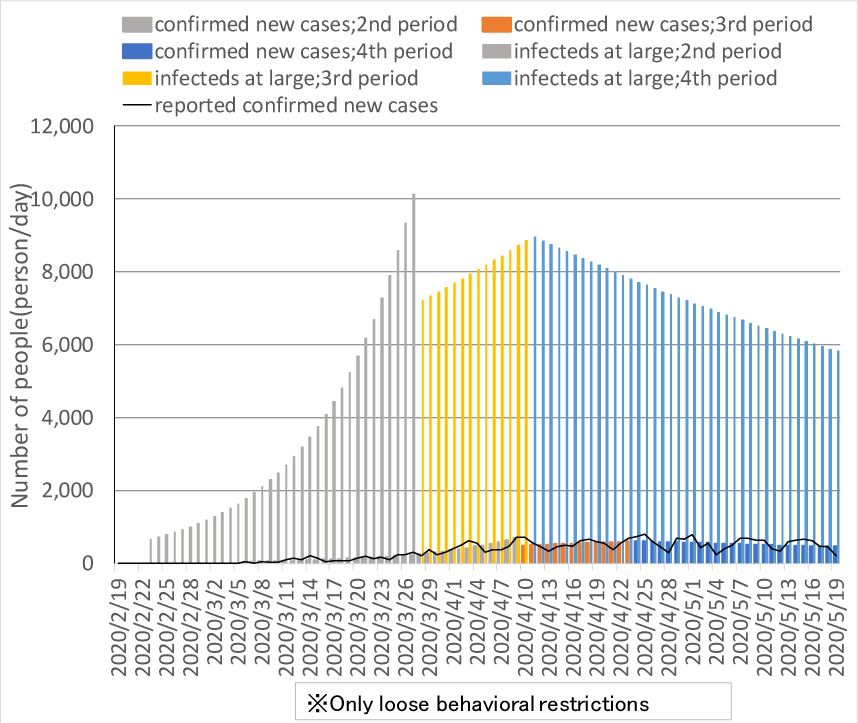
Estimation of the number of infecteds at large and confirmed new cases (Sweden)

Sweden tends to have a very low rate of the quarantine rate of infecteds at large and a very high rate of the fraction of infecteds at large. This may reflect the herd immunity policy.

## 4. Comparison of measures in each country

We divide infection control measures into the active group (λ ≤ 0.1), the average group (−0.1<λ<-0.02), and the passive group (−0.02≤ λ <-0) according to the magnitude of the decay (or growth) rate of infected at large (λ) in each country and compare them. The right direction of Figure 22 represents the level of testing/quarantine systems, and the downward direction represents the magnitude of the effectiveness of social distancing (+ early quarantine effect).

**Figure 22:**
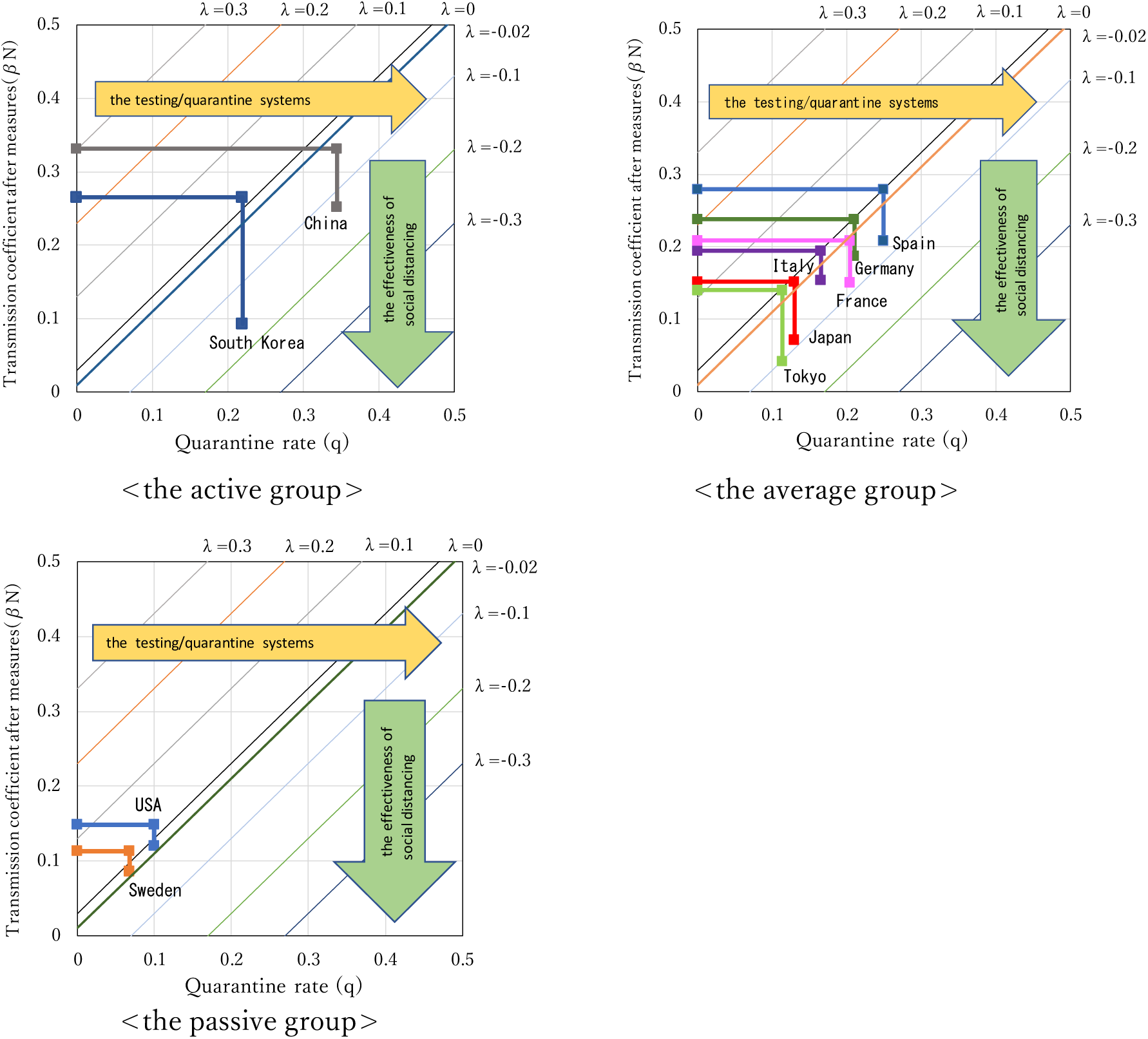
Comparison of measures in each country. Note: The effectiveness of social distancing for Japan and Tokyo are quite high (53% and 70%, respectively), but the magnitude of effectiveness of social distancing is small due to the small original transmission coefficient values.

Figure 22, which shows the comparison of measures in each country, reveals the following.

1. China and South Korea are the countries with the best testing/ quarantine systems.
2. South Korea is the country with the highest degree of effectiveness of social distancing (+ early quarantine effect), and the transmission coefficient after measures has decreased due to early quarantine through thorough testing.
3. The United States and Sweden are the only countries with a poor testing/quarantine system and a low effectiveness of social distancing. Sweden, which has a herd immunity policy, has the lowest rates for both parameters.

From the above, it can be said that the measures taken by South Korea, which did not have a lockdown system and adopted a policy of early detection and quarantine of positive cases through PCR testing, were effective.

## 5. Discussion

The following is a summary of the characteristics of the new corona infection trend in Japan.

### (1) Number of tests performed and Number of positive cases

The number of tests conducted in Japan is extremely low, at 214 per 100,000 people, compared to 1/7 in South Korea, 1/10 in France and Sweden, 1/20 in Germany and the US, 1/26 in Italy and 1/35 in Spain. The number of positive cases per 100,000 people is 13, which is very low in relation to the number of tests performed. As shown in Figure 23, which shows the relationship between the number of tests performed and the number of positive cases, the number of positive cases in Japan is extremely low compared to other countries.

**Fig.23.**
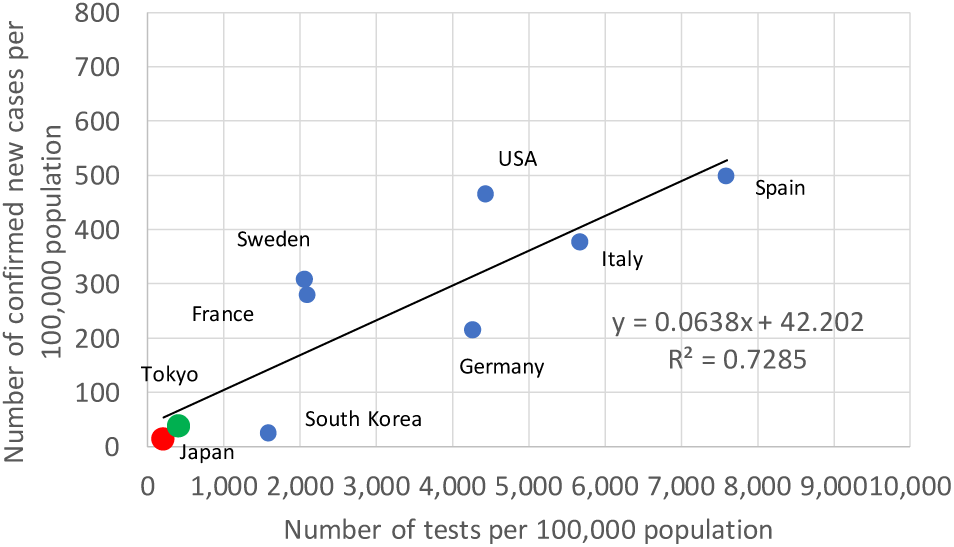
Number of tests per 100,000 population and Number of confirmed new cases per 100,000 population.

### (2) Quarantine rates of infecteds at large and Transmission coefficients

The quarantine rate of of infecteds at large in Japan is 0.130. The coefficient is 20% to 60% lower than that of other countries (0.165-0.345) except for the U. S. (0.100) and Sweden (0.0673). The transmission coefficient is 0.152. It is 20% to 50% lower than that of other countries (0.195 to 0.332) except for the U.S. (0.148) and Sweden (0.113). Figure 24 shows the trends for the quarantine rate of infecteds at large and the transmission coefficient in each country, and you can see that there is a very high correlation between the two factors.

**Fig.24.**
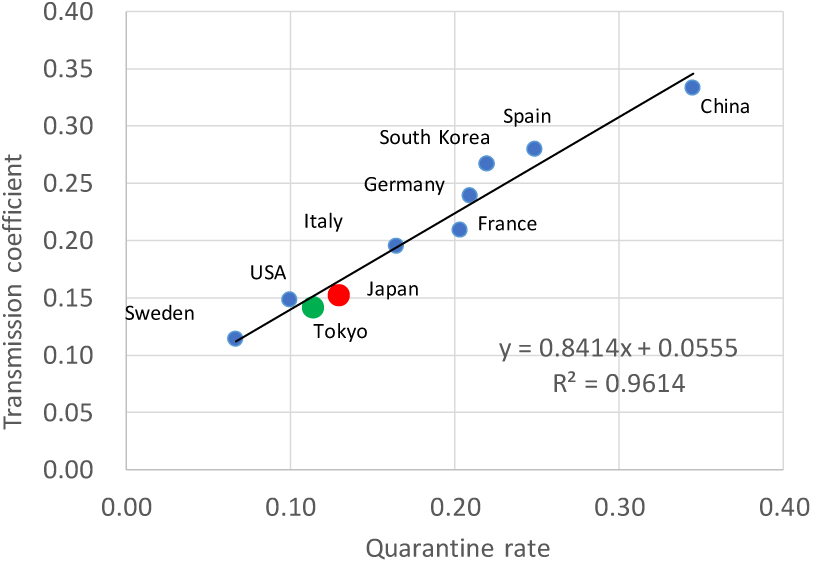
Quarantine rate and Transmission coefficient.

### (3) Quarantine rate and Effective decay (or growth) rate of infected at large

Figure 25 shows the relationship between the quarantine rate and the effective decay (or growth) rate of infected at large and the smaller the quarantine rate, the more the effective decay (or growth) rate of infected at large tends to be large. In the case of Japan, the effective decay (or growth) rate of infected at large was 8.52, following Sweden’s 14.1 and the U.S.’s 9.59.

**Fig.25.**
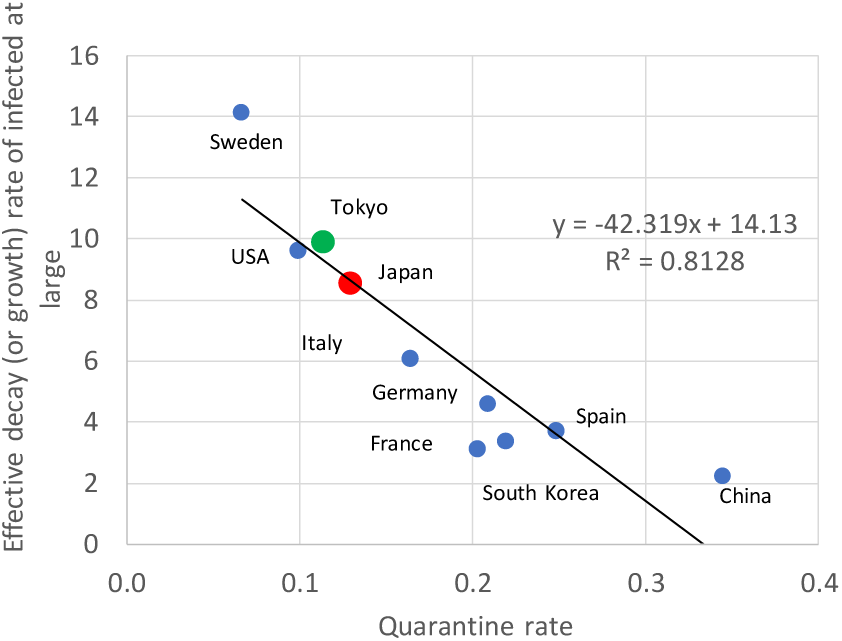
Quarantine rate and Effective decay (or growth) r ate of infected at large.

### (4) Number of positive cases and Fraction of infecteds at large in the population

Figure 26 shows the relationship between the number of positive cases per 100,000 and the fraction of infecteds at large in the population, and you can see that when the number of positive cases is low, the fraction of infecteds at large tends to be small. In Japan, the fraction of infecteds at large is 0.109%, which ranks third after China and South Korea. On the other hand, in Sweden and the U. S., where the number of positive cases per 100,000 people is high, the fraction of infecteds at large is 4.3% to 4.4%, and in Spain and Italy the rate is 1.8% to 2.3%.

**Fig.26.**
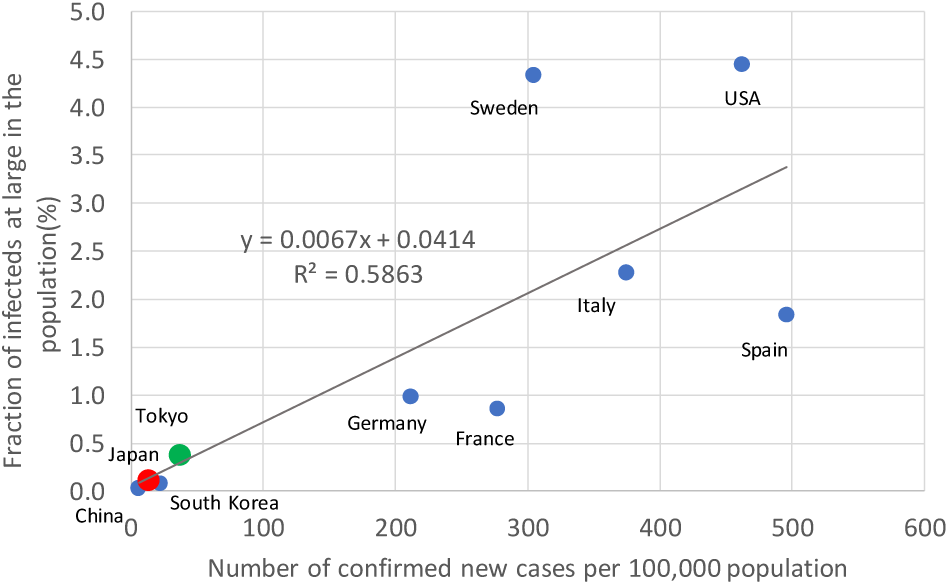
Number of confirmed new cases per 100,000 population and Fraction of infecteds at large in the population

### (5) Fraction of infecteds at large in the population and Actual antibody possession rate

The results of large-scale surveys of antibody possession rates in various countries are awaited in order to evaluate the estimates of the fraction of infecteds at large in the population based on the SIQR model. However, based on the results of surveys in Japan, the United States, Sweden, and other countries, the SIQR model has the potential to reproduce the current situation (see Table 2). In particular, the fraction of infecteds at large in the population of 0.365% in Tokyo is close to the Softbank Group’s antibody possession rate of 0.43%.

**Table 2.** Antibody test results and Fraction of infecteds at large in the population.

| Country Name | Test Group and Scope of Coverage | Testing Number(People) | Antibody retention rate | Fraction of infecteds at large in the population |
| --- | --- | --- | --- | --- |
| Japan | Kodama Group, University of Tokyo; General medical institutions in Tokyo | 500 | 0.60% | 0.109% ; Japan |
|  | SoftBank Group | 44,066 | 0.43% | 0.365% ; Tokyo |
| USA | New York State Department of Insurance | 15,000 | 12.30% | 4.43% |
|  | Santa Clara County, California. | 3,300 | 1.50% |  |
|  | Los Angeles County | 863 | 4.10% |  |
| Sweden | Stockholm |  | 7.30% | 4.32% |
| Note | • Sources for Japan, the U.S. and Sweden are shown in references 7)~10)<br>• Fraction of infecteds at large in the population are shown as validated by the SIQR model |  |  |  |

### (6) Effectiveness of social distancing and Movement reduction rate

The effectiveness of social distancing during the 4th period was estimated to be 53% in Japan as a whole and 70% in Tokyo. This is higher than in the Google Mobile Location Survey^6)^, which uses mobile device location data. In other words, the report’s data show that during the 4th period (4/19-5/19), the average decline in “retail and entertainment,” “transfer station” and “workplace” was 39% for Japan as a whole and 54% for Tokyo. In the case of Sweden, the average movement reduction rate in the same report happened to match the 24% effectiveness of social distancing in the 4th decay period, but in all other countries, the effectiveness of social distancing tended to be smaller than the mobile survey’s movement reduction rate (see Figure 27).

**Fig.27.**
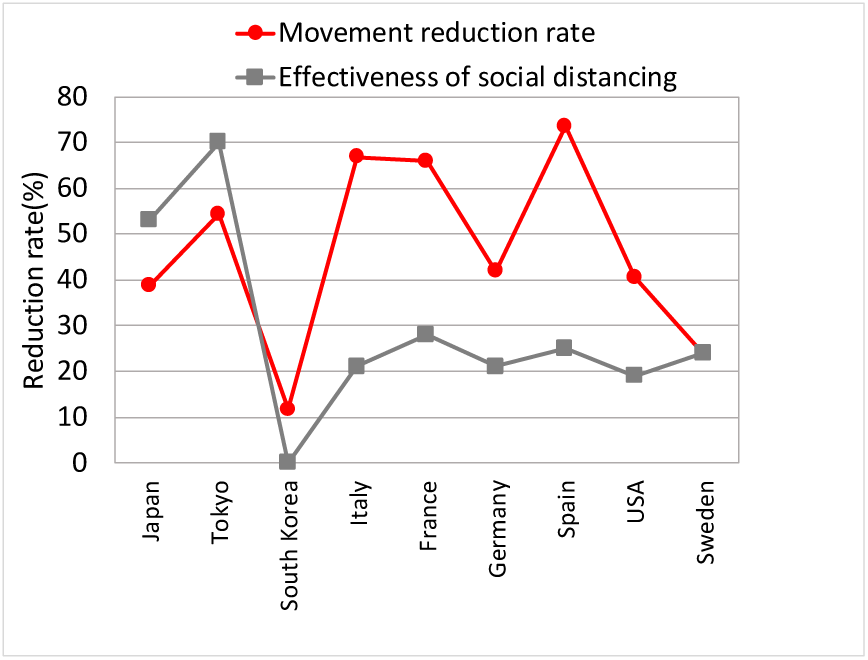
Movement reduction rate and Effectiveness of social distancing.

In the case of Japan, the level of self-restraint among its citizens is considerably higher than in the United States and Europe, but the movement reduction rate does not necessarily reflect the effectiveness of social distancing. In locked-down countries such as Italy, France, and Spain, the movement reduction rate is as high as 60-70%, while the effectiveness of social distancing is as low as 20-30%. One possible reason for this is the high fatality rate in these countries. The relationship between the fatality rate and the difference between the movement reduction rate and the effectiveness of social distancing shows a fairly high correlation (Figure 28). In other words, countries with higher fatality rates tend to have less of a lockdown effect and lower effectiveness of social distancing.

**Fig.28.**
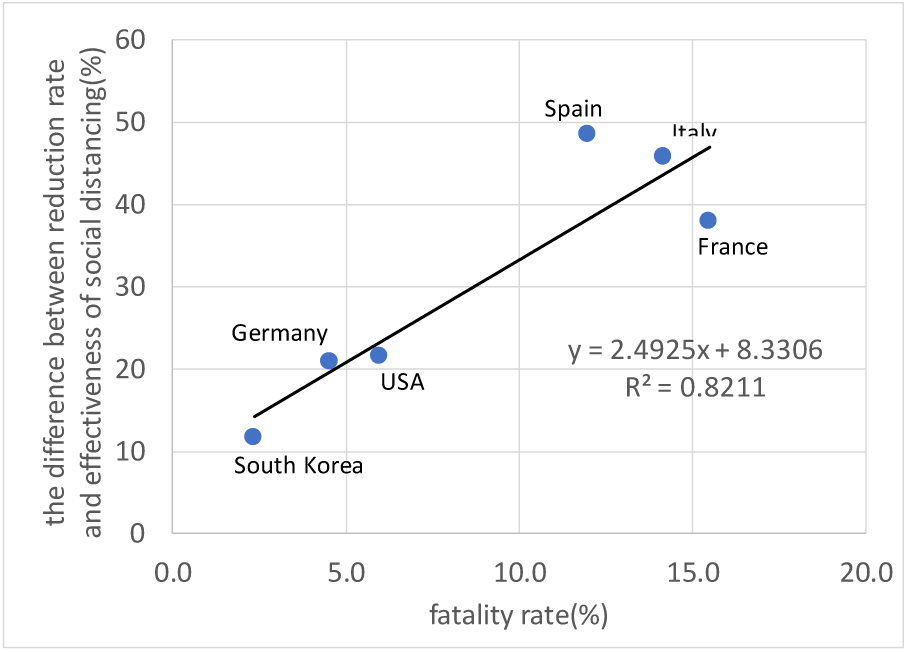
Fatality rate and Difference between the movement reduction rate and the effectiveness of social distancing.

It is important to note that the reasons for the low effectiveness of social distancing in locked-down countries are (1) the toxicity and highly infectious variants of the new coronaviruses increase the lethality rate, so that reduced movement alone may not reduce contact infection, and (2) differences in lifestyle habits, such as resistance to wearing masks, hugging, handshaking, and loud conversations, may not reduce contact infection through reduced movement alone.

### (7) The peculiarity of Japan

The number of tests conducted in Japan was extremely low, but this alone cannot explain the peculiar curve in the number of cases shown in Figure 1. Other possible reasons for Japan’s peculiarity are as follows.

1. The Ministry of Health, Labour and Welfare’s “How to Prevent a New Coronavirus” (Feb.17) announced that a “four-day fever rule” was established in which patients must have a fever of 37.5°C or higher for four days or more to be consulted. As a result of the strict implementation of the rule, the percentage of PCR tests performed in the whole country to the number of consultations with the Returnee and Contact Center was 2.9% by 11 March and 4.0% by 31 March.
2. Until the date of the decision to postpone the Olympics (March 24), it was expected that information on the number of infected people unfavorable to the hosting of the Olympic Games would be as low as possible, but verification of the SIQR model showed that the enhancement factor of quarantine rate was 0.57 during the first period of test suppression up to the date of the decision to postpone the Olympics, and that tests were suppressed by about 40% compared to the second and subsequent periods. This is the reason why the Japanese curve in Figure 1 is peculiar for the period from February 17 to March 24.
3. After the date of the decision to postpone, there is no longer a brake on the release of information on the number of infected people, but the Ministry of Health, Labour and Welfare and the National Institute of Infectious Diseases and other governmental organizations monopolized the data for unified analysis of infection data. As a result, no measures were taken to expand PCR testing by the private sector and the “four-day fever rule” continued further.
4. Verification of the SIQR model showed that as a result of test suppression, the quarantine rate of infecteds at large in Japan was 0.13, which was 20% to 60% smaller than in other countries (0.165 to 0.345) except for the United States (0.100) and Sweden (0.0673).

## Data Availability

Yes

## Acknowledgment

The author would like to thank Professor Emeritus Takashi Odagaki of Kyushu University for his SIQR model that inspired the author to complete these analyses.

## References

1) T. Odagaki, Infect. Dis. Model. {\bf 5}, 691–698 (2020).\\doi: 10.1016/j.idm.2020.08.013

2) Situation of domestic infection of the new coronavirus, Toyo Keizai ONLINE, (online), available from >https://toyokeizai.net/sp/visual/tko/covid19/>, Accessed: <2020-05-21>

3) Tokyo Metropolitan Government website for combating new coronavirus infection, (online), available from <https://stopcovid19.metro.tokyo.Ig.jp>, Accessed: <2020-05-21>

4) COVID-19 CORONAVIRUS PANDEMIC, (online), available from <https://www.worldometers.info/coronavirus/>, Accessed: <2020-05-25>

5) Coronavirus COVID-19 Global Cases by Johns Hopkins, (online), available from <https://reliefweb.int/report/world/coronavirus-covid-19-global-cases-johns-hopkins-csse>, Accessed: <2020-05-21>

6) Google LLC “Google COVID-19 Community Mobility Reports”, (online), available from < https://www.google.com/covid19/mobility/> Accessed: <2020-06-05>

7) Tatsuhiko Kodama and Takeshi Kawamura, Results of Tokyo’s antibody test (press conference), University of Tokyo Advanced Science Technology Research Center, May 15, 2020

8) Softbank News _ Preliminary antibody test results and exit strategy, <https://www.softbank.jp/sbnews/entry/20200610_01>. Accessed: <2020-06-12>

9) Masahiro Kami, Twitter May 7, 2020, Antibody possession rates and excess deaths in Japan and abroad, Erika Yamashita Web survey dates May 5–7, 2020

10) Tokyo Shimbun (morning edition), Testing for antibodies to the new corona in each country, June 7, 2020

